# Clinical epidemiology of the endoscopic, laparoscopic, and surgical resection of malignant gastric tumors in Japan, 2014-2021: a retrospective study using open data from a national claims database

**DOI:** 10.1101/2024.04.30.24305814

**Authors:** Akahito Sako, Tomoyuki Yada, Keiichi Fujiya, Ryo Nakashima, Kensuke Yoshimura, Hidekatsu Yanai, Naomi Uemura

## Abstract

**Background:** Gastric cancer is one of the most common malignancies and its incidence is high in East Asia. Several options are available for resection of malignant gastric tumors, ranging from endoscopic resection of early-stage cancer to open total gastrectomy. However, there has been a lack of nationwide data on gastric resection in Japan.

**Methods:** This observational study analyzed data from the publicly accessible National Database of Health Insurance Claims and Specific Health Checkups, which includes most national health insurance claims data in Japan. Trends in the various types of resection performed for malignant gastric tumors between 2014 and 2021, the age and sex distributions of patients undergoing these procedures, and regional disparities were investigated.

**Results:** The annual number of resections was highest in 2015 (109,000) and lowest in 2020 (90,000) after the COVID-19 pandemic. The proportion of endoscopic resections increased from 47% in 2014 to 57% in 2021 while the proportion of total gastrectomies performed during this period decreased from 17% to 10%. In 2021, 70% of patients who underwent resection were men. That year, 83.8% of all patients who underwent any type of gastric resection and 87.1% of those who underwent endoscopic submucosal dissection (ESD) were aged ≥65 years. The annual incidence of gastric resection per million general population was highest in Tottori (n=1,236) and lowest in Okinawa (n=251). The proportion of endoscopic resections was highest in Miyagi (66%) and lowest in Aichi (45%) and that of open surgery was highest in Aomori (36%) and lowest in Wakayama (5%).

**Conclusions:** Gastric malignancy is increasingly treated by endoscopic submucosal dissection rather than by open total gastrectomy. However, there are regional disparities in the resection methods used. Standardization of screening and treatment and a more even distribution of specialists are needed.

## BACKGROUND

Gastric cancer is one of the most common malignancies worldwide and has a high incidence in East Asia and South America due to their higher prevalence of *Helicobacter pylori* infection (1, 2). Prompt diagnosis of gastric cancer is important because of the higher likelihood of curative treatment in the early stages and the greater opportunity to use less invasive treatment options, such as endoscopic submucosal dissection (ESD).

In Japan, the Cancer Control Act enacted in 2007 identified the basic means of cancer control to be consistency of treatment, prevention and early diagnosis of cancer, and promotion of cancer research. In Japan, there are regional disparities in the rate of *H. pylori* infection and in the incidence and mortality of gastric cancer (3, 4). However, data on the treatment of gastric cancer according to region are limited. Detailed nationwide or large-scale data on endoscopic and surgical resection of gastric malignancy are lacking in many countries, including Japan. The National Clinical Database (NCD) contains nationwide data on most of the surgeries performed in Japan, including patient characteristics, indications for and types of surgery, and morbidity and mortality (5–9), but does not include endoscopic resection.

In this descriptive study, we investigated the clinical epidemiology of endoscopic and surgical resection of malignant gastric tumors in Japan by analyzing the nearly complete enumeration in the dataset held by a national health insurance claims database.

## METHODS

This retrospective cohort study analyzed open data from the National Database of Health Insurance Claims and Specific Health Checkups (NDB) of Japan. Japan has a population of 126 million and a universal health care system. The Ministry of Health, Labour and Welfare has used the NDB to collect almost all national health insurance claims data for both inpatients and outpatients since 2009. The NDB contains data on patient sex, age, diagnosis, examinations, prescriptions, and surgeries. NDB Open Data, an anonymized dataset extracted from the NDB and made publicly available on the NDB website, does not provide individual data; it provides aggregate data, including annual numbers of surgeries and procedures performed in Japan stratified by sex, 5-year age intervals, inpatient/outpatient status, month, and geographic location of clinics or hospitals in Japan’s 47 prefectures (from Hokkaido in the north to Okinawa in the south) (10). Age distribution by sex is also available, but the age distribution within each prefecture is not. No diagnoses are available. In Japan, the fiscal year (FY) runs from April 1 to March 31, and as of December 2023, data are available for FY2014 to FY2021. We analyzed data for this entire period with a focus on FY2021. For the denominator, we used the population of Japan stratified by age, sex, and prefecture, as reported by the Statistics Bureau of Japan (11).

Data from NDB Open Data are anonymized, so there are some missing values in the data. Data are available in the form of an Excel spreadsheet for the total annual number of surgeries and distribution by sex, 5-year age interval, and prefecture. However, if the number of surgeries in a specific cell is low, the data are masked. The cut-off value for masking is 10. For example, if only 8 ESDs were performed for men aged 20-24 years in 2021, this figure would be masked.

Many studies have examined data from the NDB (12, 13) and NDB Open Data (14–16). The institutional review board of the National Center for Global Health and Medicine reviewed and approved the study protocol. The need for informed consent was waived because the data in NDB Open Data are anonymized and publicly available.

### Endoscopic, laparoscopic, and surgical resection of malignant gastric tumors

For the purposes of the Japanese health reimbursement system, endoscopic resection for malignant gastric tumors is divided into polypectomy, endoscopic mucosal resection (EMR), and ESD for early-stage malignant tumors of the stomach and duodenum. ESD for the stomach and ESD for the duodenum have been differentiated since 2020. We did not include endoscopic resection for non-malignant tumors. Local gastrectomy is divided into open, laparoscopic, and laparoscopic and endoscopic collaborative surgery (LECS). LECS for the duodenum has been differentiated from LECS for the stomach since 2020. Extent of resection was divided into 6 groups, namely, EMR and polypectomy, ESD, local gastrectomy, distal gastrectomy, proximal gastrectomy, and total gastrectomy. Surgical procedures were categorized as LECS, endoscopic, robotic-assisted, laparoscopic, and open surgery. Robotic-assisted gastrectomy was first approved for Advanced Medical Technology in 2014 and first covered by public health insurance and included in the NDB in 2018. Gastrectomy for non-malignant disease was excluded.

## RESULTS

### Annual numbers and types of resection for malignant gastric tumors

Between 2014 and 2019, the annual number of resections performed for malignant gastric tumors ranged from 103,000 to 109,000 (Table 1). In 2020, this number decreased to 90,000, which coincided with a decrease in the number of upper gastrointestinal endoscopies performed for examination purposes. The proportion of endoscopic resections increased from 46.9% in 2014 to 56.5% in 2021. ESD accounted for 92-93% of endoscopic resections. The proportion of laparoscopic surgeries increased from 19.8% in 2014 to 24.4% in 2021. Robot-assisted surgery accounted for 5.5% of all laparoscopic surgeries performed in 2018 and 16.8% of those performed in 2021. The proportion of total gastrectomies performed decreased from 16.5% in 2014 to 10.0% in 2021.

**Table 1.**
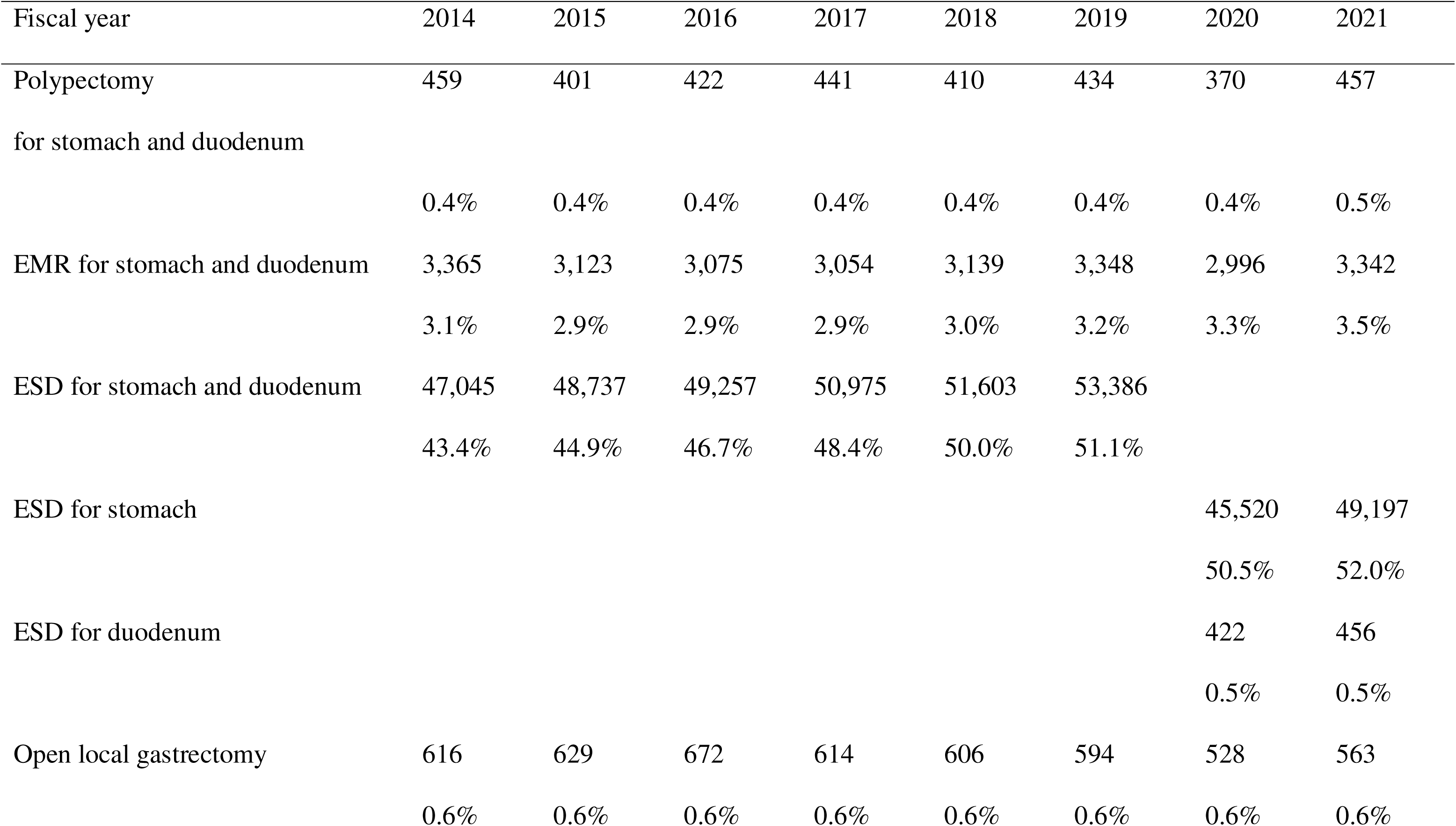
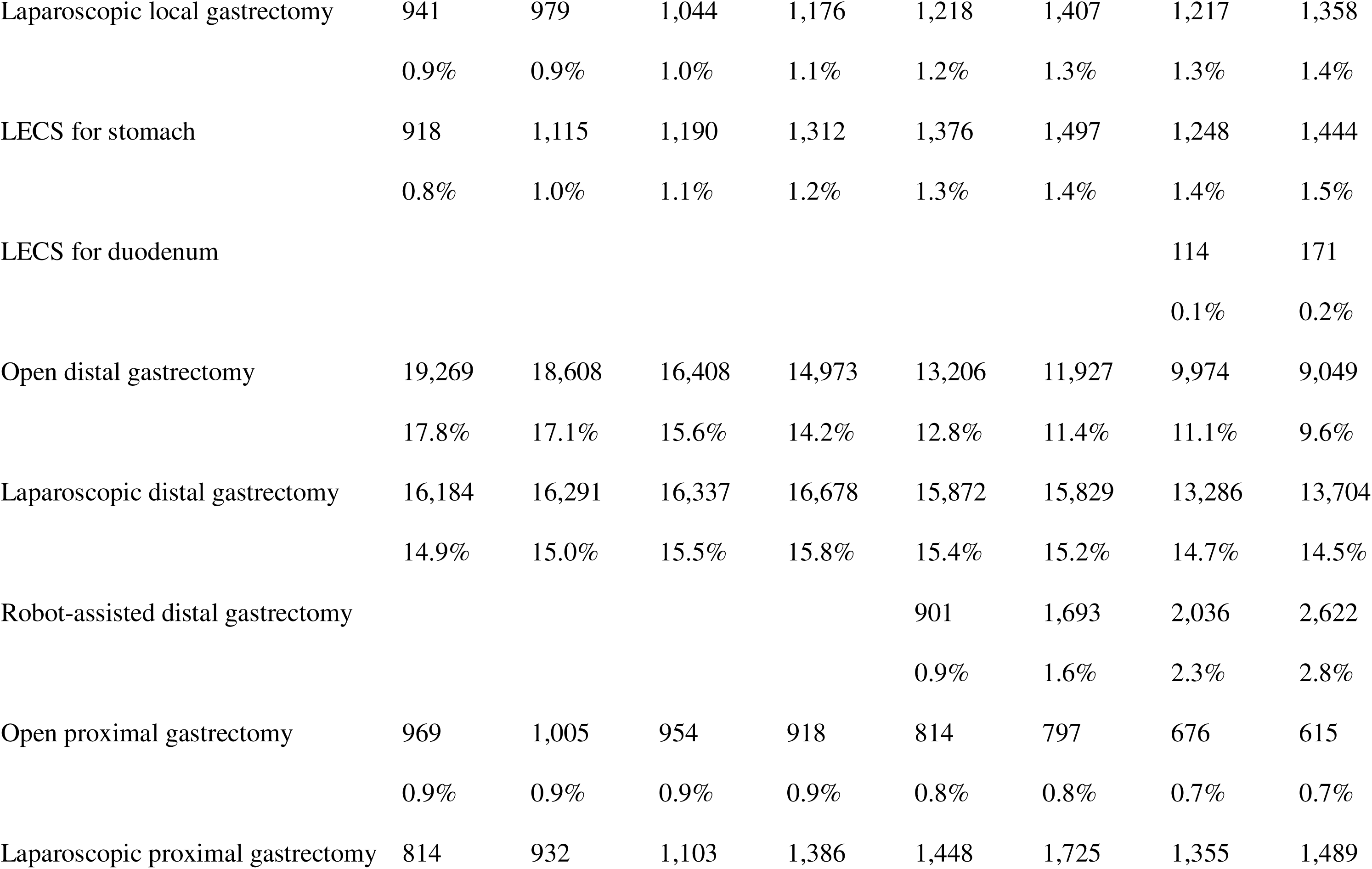
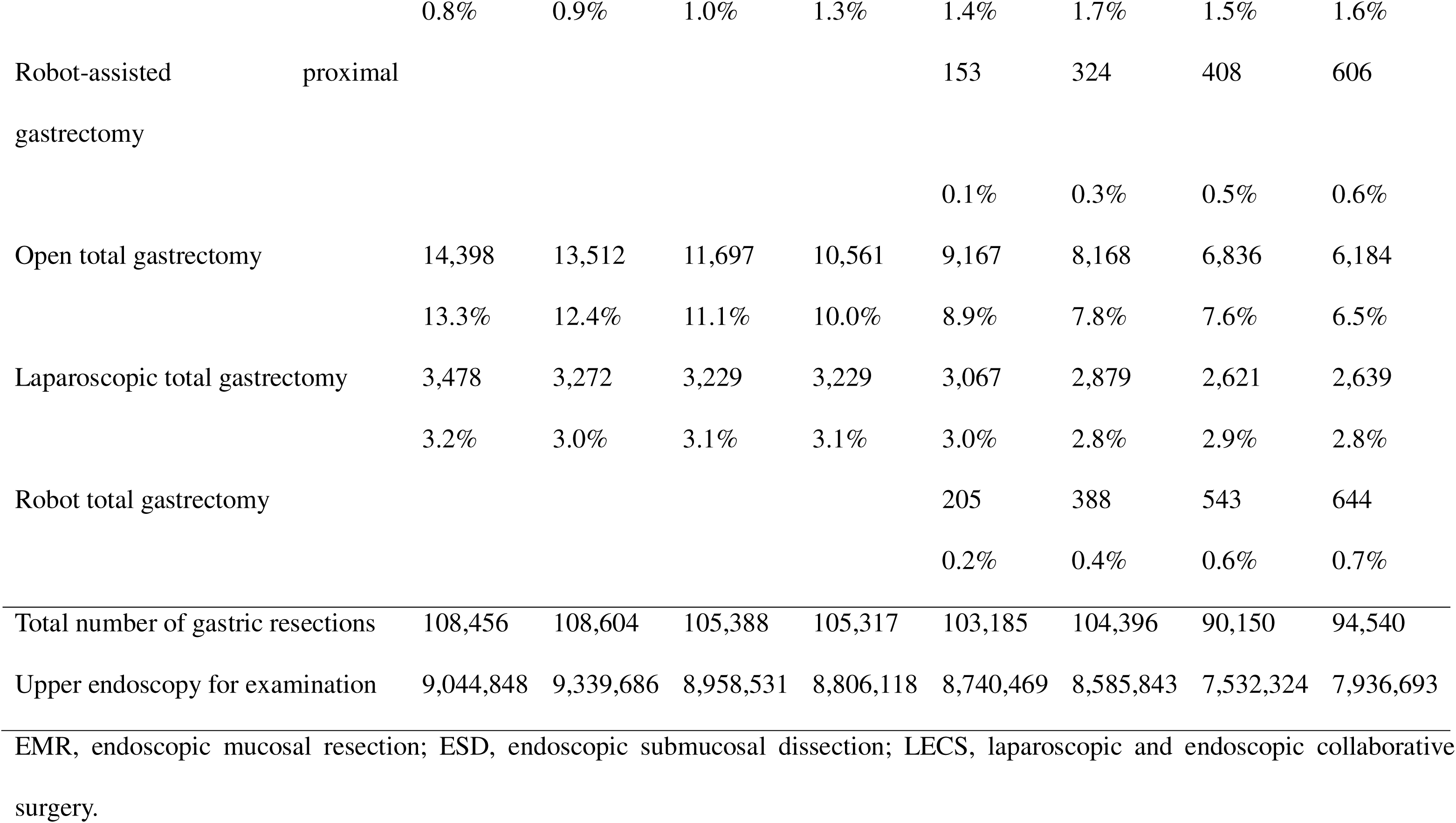
Annual numbers of gastric resections and upper endoscopies performed between 2014 and 2021.

### Age and sex distribution in 2021

In 2021, 69.7% of patients who underwent resection of malignant gastric tumors were men. Although men accounted for 60-80% of most types of resection, similar numbers of men and women underwent local gastrectomy, including LECS.

Patients aged ≥65 years and ≥75 years respectively accounted for 83.8% and 48.7% of all types of gastric resection and 87.1% and 52.0% of ESDs performed for malignant gastric tumors.

The incidence of gastric resection was highest in patients aged 80-84 years irrespective of sex (Fig 1). The annual incidence of gastric resection per million general population aged 80-84 years was 4,819 for men and 1,599 for women.

**Figure 1:**
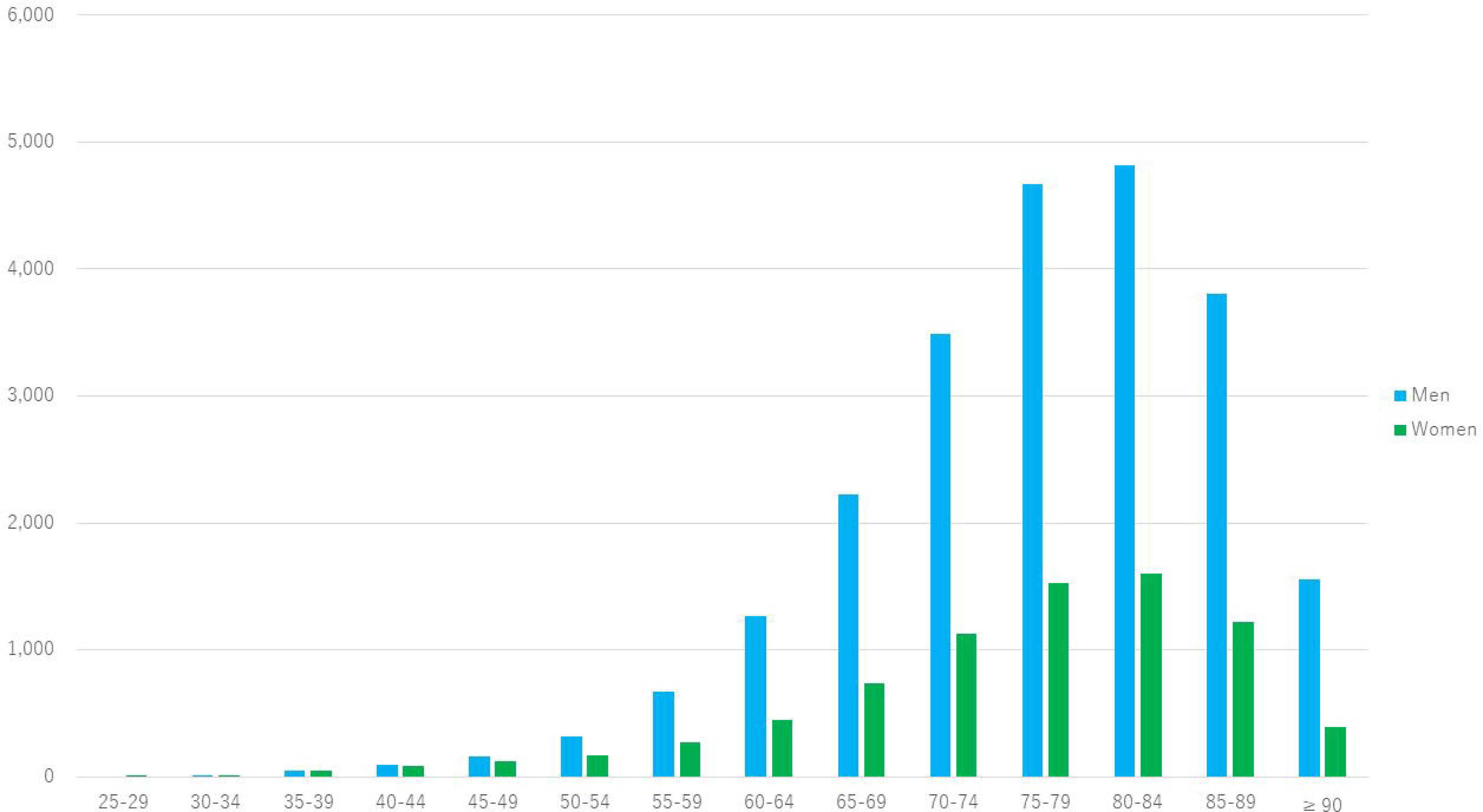
Gastric resections performed per million general population in 2021 by age and sex.

In terms of extent of resection, the proportions of endoscopic resection, LECS and local gastrectomy, distal gastrectomy, proximal gastrectomy, and total gastrectomy were 56.8%, 3.6%, 26.9%, 2.8%, and 10.0%, respectively (Fig 2). LECS and local gastrectomy were more common in younger patients. The older the patient, the more likely they were to undergo endoscopic resection and the lower the likelihood of total gastrectomy.

**Figure 2:**
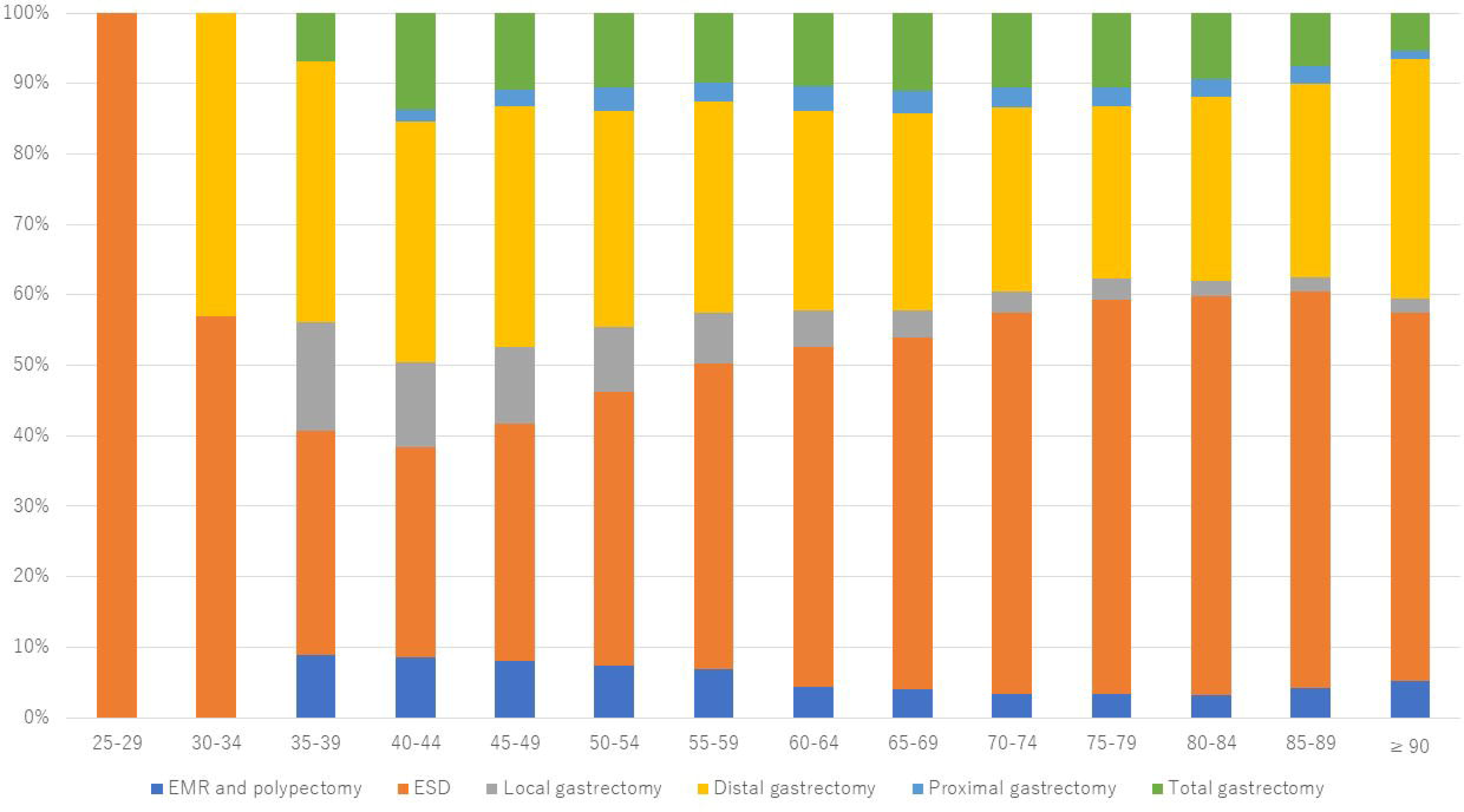
Extent of resections performed in 2021 by age. EMR, endoscopic mucosal resection; ESD, endoscopic submucosal dissection.

In regard to the resection procedures performed, the respective proportions of endoscopic resection, LECS, robot-assisted surgery, laparoscopic surgery, and open surgery were 56.8%, 1.6%, 4.0%, 20.3%, and 17.3% (Fig 3). Open surgery was performed predominantly in older patients and laparoscopic and robot surgery in relatively young patients.

**Figure 3:**
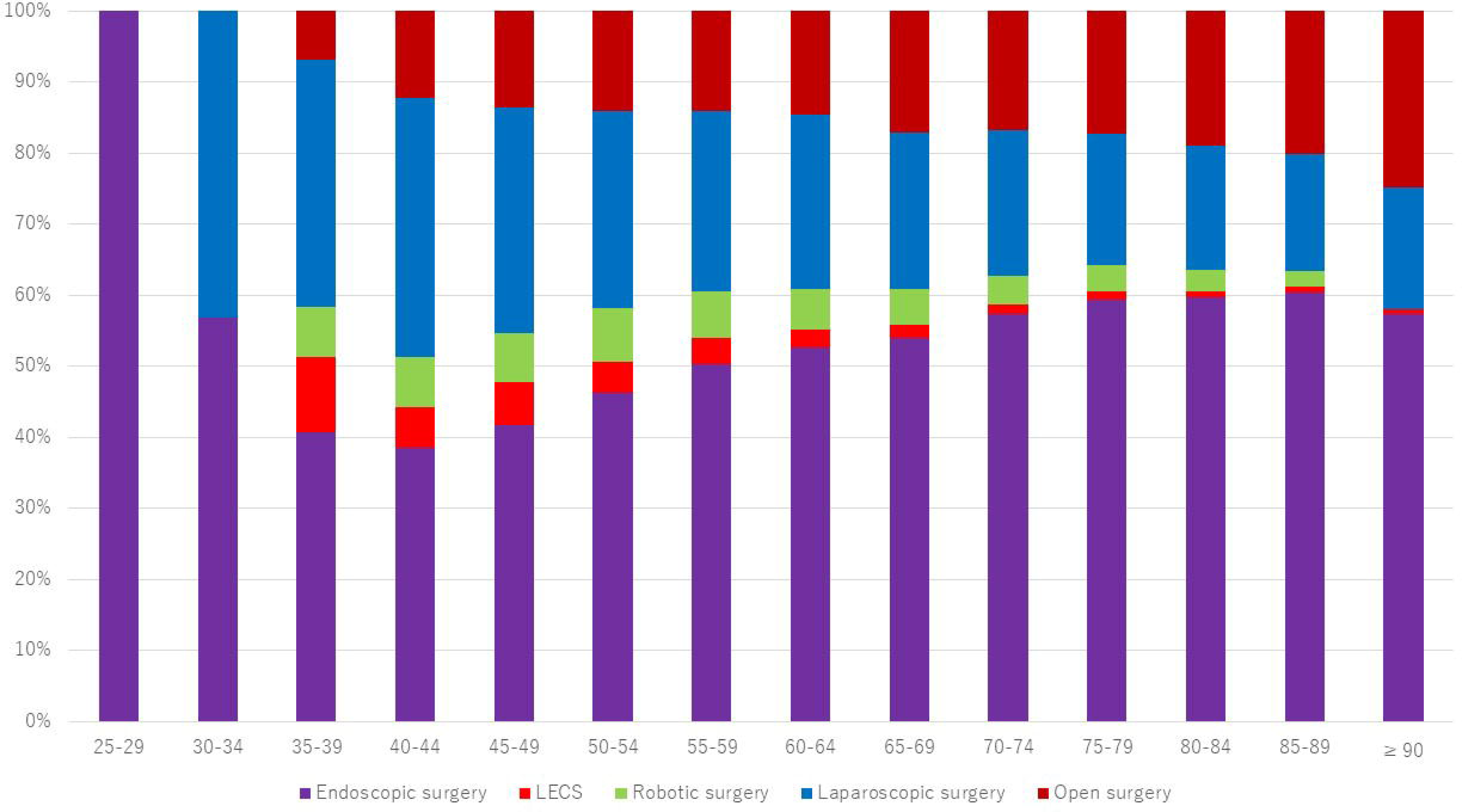
Types of surgical resection performed in 2021 by age.

### Geographic differences by prefecture in 2021

In 2021, the incidence of gastric resection per million general population was 753 in Japan. It was highest in Tottori (1,236), followed by Niigata (1,225) and Yamagata (1,207), and lowest in Okinawa (251) followed by Miyazaki (528) and Aichi (529) (Fig 4).

**Figure 4:**
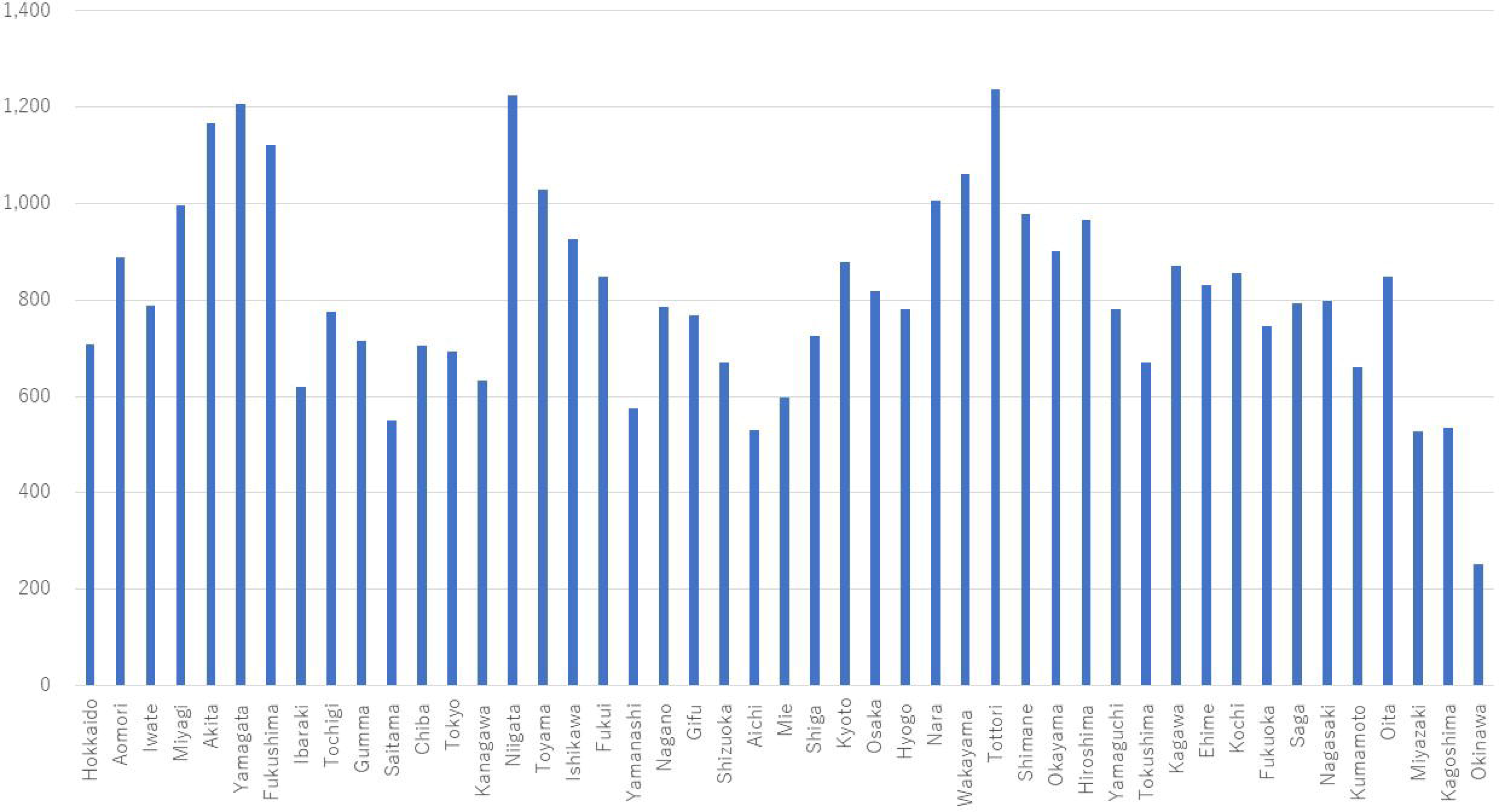
Gastric resections performed per million general population in 2021 by prefecture.

In terms of extent of resection, the proportion of patients who underwent total gastrectomy was highest in Miyazaki (16.3%), followed by Akita and Aomori (14.6%), and lowest in Kagawa and Fukui (5.7%), followed by Kyoto (6.3%) (Fig 5). The proportion of patients who underwent endoscopic resection was highest in Miyagi (66.2%), followed by Shimane (65.3%) and Kagawa (64.4), and lowest in Aichi (44.7%), followed by Okinawa (45.7%) and Yamaguchi (48.3%).

**Figure 5:**
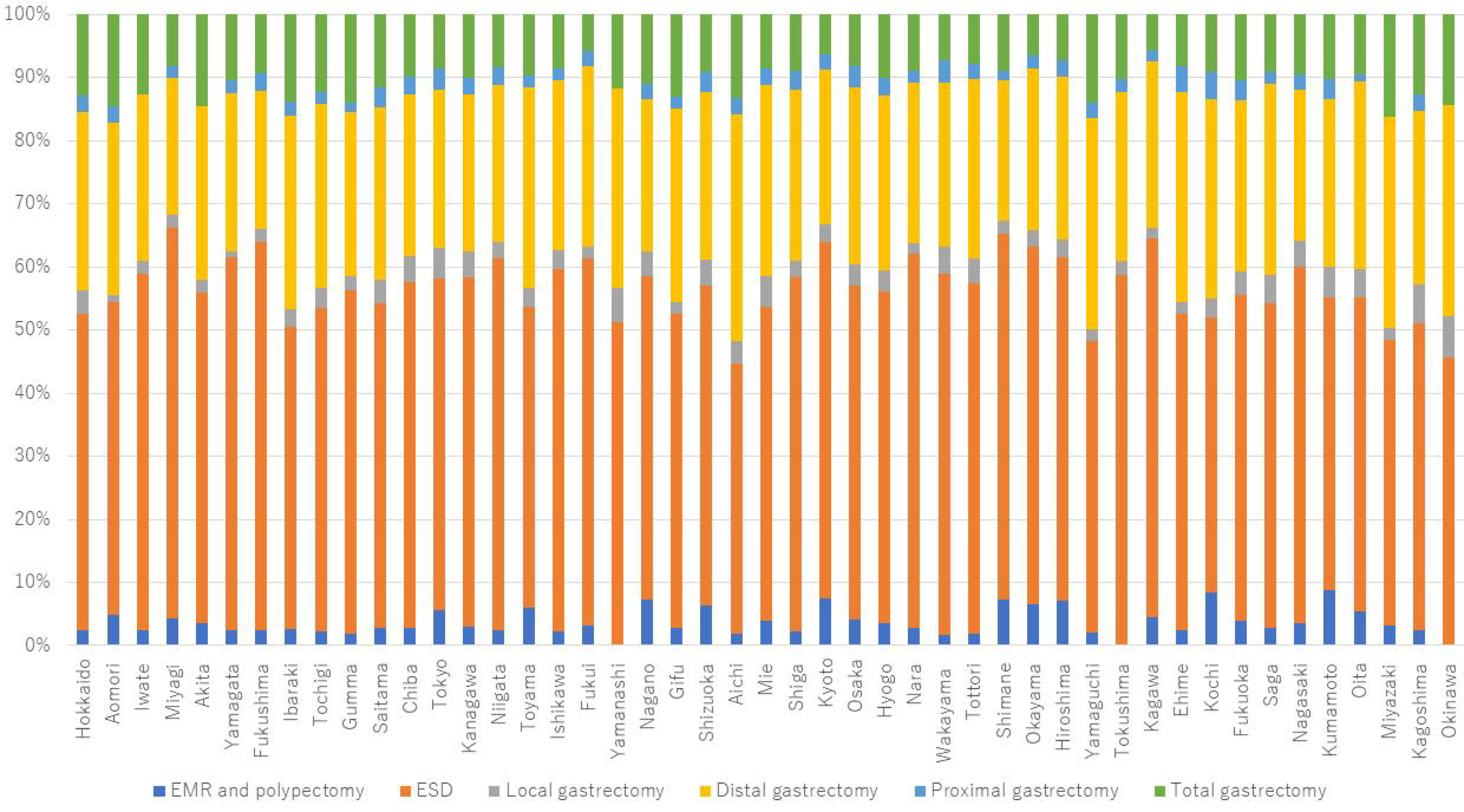
Extent of resections performed in 2021 by prefecture. EMR, endoscopic mucosal resection; ESD, endoscopic submucosal dissection.

With regard to type of resection, open surgery was most common in Aomori (36.0%), followed by Miyazaki (29.3%) and Ibaraki (28.8%), and lowest in Wakayama (5.3%), followed by Saga (7.2%) and Yamanashi (8.4%) (Fig 6). Robot-assisted surgery was performed most often in Aichi (10.9%), followed by Wakayama (10.1%) and Kochi (8.9%), and 0% in 9 prefectures.

**Figure 6:**
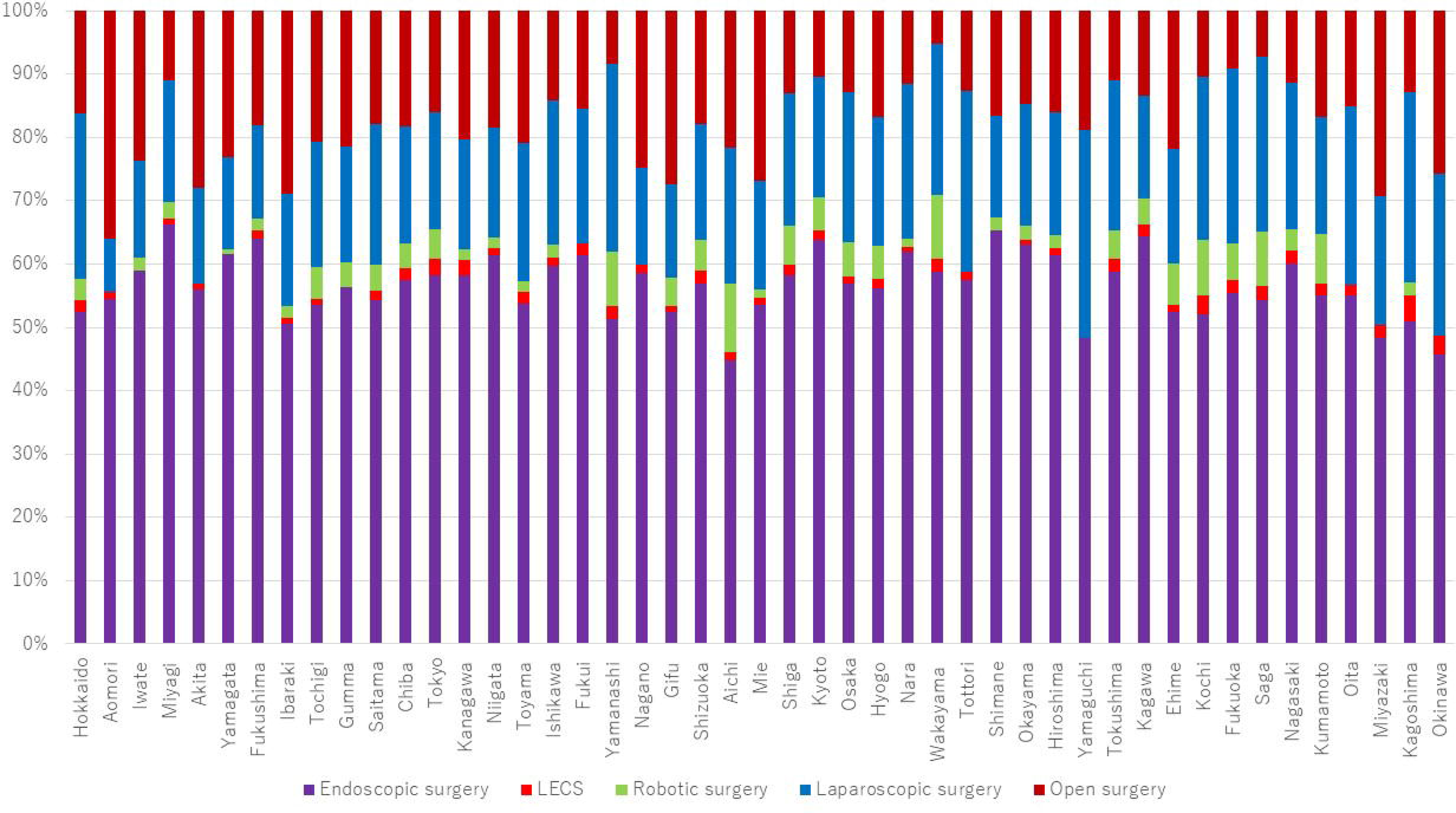
Types of surgical resection performed in 2021 by prefecture.

## DISCUSSION

This nationwide retrospective study investigated trends in age and sex distribution as well as regional disparities in resection of malignant gastric tumors in Japan during the 8 years between 2014 and 2021. About 100,000 resections were performed annually; 48.7% were performed in patients aged ≥75 years and 69.7% were men. Large regional disparities in the incidence and type of gastric resection were identified.

The NCD collects surgical data in Japan. The majority of surgical procedures for malignant gastric tumors, including laparoscopic surgery, are included in this database. Several studies of gastric cancer surgery have used the NCD to obtain information on intra-abdominal infections after surgery, long-term survival, surgery-related mortality and comparison between laparoscopic and robot-assisted surgery (5–9). However, the NCD does not contain data on endoscopic surgeries, such as ESD. The Japan Gastroenterological Endoscopy Society started the Japan Endoscopy Database (JED) in 2015 to collect information on endoscopic examinations and treatments (17). However, its coverage is insufficient in terms of the number of participating facilities and endoscopic procedures performed. Moreover, there are still no published studies of gastric cancer based on data from the JED. Although a national cancer registry system was established in Japan in 2016 (18), it is still not widely used and there are currently no published papers concerning gastric cancer that have used data from this registry. Other administrative claims databases include the NDB (12) and the Diagnosis Procedure Combination database (19, 20). These databases, unlike the NCD and JED, need less workload for data registration and allow investigation of both surgical and endoscopic procedures. For better clinical practice and evidence-based policy-making, it is important to contribute to these databases and use the data effectively.

Between FY2014 and FY2019, there was a gradual decrease in both the number of malignant gastric tumor resections and the number of upper gastrointestinal endoscopies performed, possibly due to *H. pylori* eradication therapy for peptic ulcer becoming covered by health insurance in FY2002 and *H. pylori*-infected gastritis being included in FY2013 (21, 22). In FY2020, the number of gastric malignant tumor resections and upper gastrointestinal endoscopies decreased to 86.4% and 87.7%, respectively, compared with FY2019. This decline is attributed to fewer cancer screenings and endoscopic examinations and delays in treatment caused by the COVID-19 pandemic (23, 24); however, these showed recovery in FY2021. Over time, the numbers of total gastrectomies and open gastrectomies have decreased, with those of open total gastrectomies and open distal gastrectomies having decreased by about half. In the meantime, the frequency of endoscopic resection has increased. Moreover, in 2021, robot-assisted surgery accounted for 19.6% of laparoscopic total gastrectomies and 16.1% of distal gastrectomies. The Japanese guidelines for treatment of gastric cancer expanded the indications for ESD in 2018 and 2021 and added a recommendation for robot-assisted surgery in 2021 (25). These changes in the clinical guidelines and population aging are possible reasons for the increase in cases that are treated less invasively and the reduction in open and total gastrectomies performed. However, data from NDB Open Data, which do not include detailed information, cannot explain the changes in treatment that have occurred over time.

More men underwent gastric resection during our study period, which is consistent with the fact that gastric cancer is more common in men and the elderly (2). The relatively balanced sex distribution and predominance of local gastrectomy, including LECS, in non-elderly patients is thought to reflect the characteristics of gastrointestinal stromal tumors (GISTs) (26), which is a principal target for such procedures. The majority of patients who need endoscopic or surgical resection for gastric malignancy are elderly. Therefore, there is a need for treatment that is less invasive than open and total gastrectomy and for further research on the risk of complications.

The present study is the first to show regional disparities in the choice of resection procedures for malignant gastric tumors. Previous studies using NDB Open Data have shown regional disparities in clinical practice for nephrectomy and nephroureterectomy (16) as well as prescriptions for chronic kidney disease and influenza (14, 15).

Differences in age distribution, prevalence and strains of *H. pylori*, and in the rate of participation in cancer screening lead to differences in the incidence of cancer and the clinical stage at diagnosis. An uneven distribution of specialists and facilities, lack of standardized treatment, and inadequate provision of clinical training might explain these variations (27, 28). The establishment and dissemination of clinical guidelines is an important factor in efforts to reduce regional disparities and standardize clinical treatment of gastric cancer (29, 30). The Japanese guidelines for treatment of gastric cancer were first published in 2001 and the guidelines for ESD and EMR for early gastric cancer in 2014, and both have been updated several times (25, 31). However, standardization of treatment and reduction of regional disparities by dissemination of clinical guidelines remain suboptimal.

This study has several limitations. First, patient characteristics other than age, sex, prefecture, inpatient/outpatient status, and outcome data were not available. Second, NDB Open Data contains aggregated data but not individual data. Therefore, it is not known how many patients underwent more than one endoscopic resection or how many underwent surgery after incomplete endoscopic resection. Although these numbers must have been similar to the number of gastric resections, we could not precisely calculate them. Third, as mentioned in the Methods section, some values were missing from the dataset. For example, Figure 2 shows that all of the resections performed in the 25-29-year age group was ESDs. The only data available for this age group were for 10 ESDs performed in women, and it is unknown whether other types of resection were performed in fewer than 10 cases. However, values were missing for only 4.9% of prefectures and for 0.6% in terms of information on age and sex. Therefore, the influence of missing values is likely to be small. Fourth, no clinical or pathological diagnoses were available. Malignant gastric tumors might include gastric lymphoma and GIST, and reimbursement for local gastrectomy is not limited to malignant tumors. Before 2020, ESD and LECS might have included malignant duodenal tumors. However, in this study, most gastric resections are likely to have been performed for gastric cancer because our data are similar to the number of surgeries for gastric cancer in the NCD. Fifth, there has been some inconsistency in the diagnosis of early gastric cancer and adenoma among pathologists in East Asia and their counterparts in North America and Europe (32). Moreover, the reimbursement price for ESD in patients with gastric cancer is higher than that for EMR in those with gastric cancer and endoscopic resection for non-malignant gastric tumors. Gastric adenoma can be included in ESD for gastric malignancy in view of the differences in pathological diagnosis and upcoding for reimbursement.

Despite these limitations, our study sheds light on the clinical epidemiology of patients undergoing resection of malignancy gastric tumors in terms of age and sex distribution, trends over time, and regional disparities. This is the first Japanese study to investigate both endoscopic and surgical resection using nationwide data. We believe that our findings will be useful for improvement of clinical practice and development of health care policy. Use of ESD and robot-assisted surgery has been increasing and will continue to do so in parallel with population aging and development and dissemination of novel techniques. Standardization of treatment by clinical guidelines and a more even distribution of specialists will be key to improvement of the present regional disparities in the treatment of gastric cancer across Japan. The JED database should be expanded to include nationwide clinical data on endoscopic surgery.

## Abbreviations

NDB: National Database of Health Insurance Claims and Specific Health Checkups
FY: fiscal year
EMR: endoscopic mucosal resection
ESD: endoscopic submucosal dissection
LECS: laparoscopic and endoscopic collaborative surgery
JED: Japan Endoscopy Database
GIST: gastrointestinal stromal tumor
NCD: National Clinical Database

## Declarations

### Consent for publication

Not applicable.

### Availability of data and materials

The datasets used and/or analyzed in this study are publicly available.

### Authors’ contributions

AS is the guarantor of this work, had full access to all data in the study, and takes responsibility for the integrity of the data and the accuracy of the data analysis. All authors approved the final version of the manuscript.

Study concept and design: AS

Acquisition, analysis, or interpretation of data: AS

Drafting of the manuscript: AS

Critical revision of the manuscript for important intellectual content: AS, TY, KF, RN, NU

Administrative, technical, or material support: AS, HY

Study supervision: KY, HY, NU

## Data Availability

All data produced in the present study are available upon reasonable request to the authors

## Acknowledgements

Not applicable

## REFERENCES

1. Wong MCS, Huang J, Chan PSF, Choi P, Lao XQ, Chan SM, et al. Global Incidence and Mortality of Gastric Cancer, 1980-2018. JAMA Netw Open. 2021;4(7):e2118457.

2. Luo G, Zhang Y, Guo P, Wang L, Huang Y, Li K. Global patterns and trends in stomach cancer incidence: Age, period and birth cohort analysis. Int J Cancer. 2017;141(7):1333–44.

3. National Cancer Center. Cancer Statistics in Japan 2024 [Available from: https://ganjoho.jp/reg_stat/statistics/stat/cancer/5_stomach.html.

4. Oki S, Takeda T, Hojo M, Uchida R, Suzuki N, Abe D, et al. Comparative Study of Helicobacter pylori-Infected Gastritis in Okinawa and Tokyo Based on the Kyoto Classification of Gastritis. J Clin Med. 2022;11(19).

5. Suda K, Yamamoto H, Nishigori T, Obama K, Yoda Y, Hikage M, et al. Safe implementation of robotic gastrectomy for gastric cancer under the requirements for universal health insurance coverage: a retrospective cohort study using a nationwide registry database in Japan. Gastric Cancer. 2022;25(2):438–49.

6. Iwatsuki M, Yamamoto H, Miyata H, Kakeji Y, Yoshida K, Konno H, et al. Association of surgeon and hospital volume with postoperative mortality after total gastrectomy for gastric cancer: data from 71,307 Japanese patients collected from a nationwide web-based data entry system. Gastric Cancer. 2021;24(2):526–34.

7. Suzuki S, Takahashi A, Ishikawa T, Akazawa K, Katai H, Isobe Y, et al. Surgically treated gastric cancer in Japan: 2011 annual report of the national clinical database gastric cancer registry. Gastric Cancer. 2021;24(3):545–66.

8. Kakeji Y, Takahashi A, Hasegawa H, Ueno H, Eguchi S, Endo I, et al. Surgical outcomes in gastroenterological surgery in Japan: Report of the National Clinical Database 2011-2018. Ann Gastroenterol Surg. 2020;4(3):250–74.

9. Fujiya K, Kumamaru H, Fujiwara Y, Miyata H, Tsuburaya A, Kodera Y, et al. Preoperative risk factors for postoperative intra-abdominal infectious complication after gastrectomy for gastric cancer using a Japanese web-based nationwide database. Gastric Cancer. 2021;24(1):205–13.

10. Ministry of Health, Labour and Welfare. NDB Open Data 2024 [Available from: https://www.mhlw.go.jp/stf/seisakunitsuite/bunya/0000177182.html.

11. Statistics Bureau of Japan. Portal Site of Official Statistics of Japan 2024 [Available from: https://www.e-stat.go.jp/en.

12. Ishimaru M, Matsui H, Ono S, Hagiwara Y, Morita K, Yasunaga H. Preoperative oral care and effect on postoperative complications after major cancer surgery. Br J Surg. 2018;105(12):1688–96.

13. Sugiyama T, Imai K, Ihana-Sugiyama N, Tanaka H, Yanagisawa-Sugita A, Sasako T, et al. Variation in process quality measures of diabetes care by region and institution in Japan during 2015-2016: An observational study of nationwide claims data. Diabetes research and clinical practice. 2019;155:107750.

14. Sako A, Gu Y, Masui Y, Yoshimura K, Yanai H, Ohmagari N. Prescription of anti-influenza drugs in Japan, 2014-2020: A retrospective study using open data from the national claims database. PloS one. 2023;18(10):e0291673.

15. Inoue R, Nishi H, Tanaka T, Nangaku M. Regional variance in patterns of prescriptions for chronic kidney disease in Japan. Clin Exp Nephrol. 2019;23(6):859–64.

16. Sugihara T, Yasunaga H, Matsui H, Kamei J, Fujimura T, Kume H. Regional clinical practice variation in urology: Usage example of the Open Data of the National Database of Health Insurance Claims and Specific Health Checkups of Japan. Int J Urol. 2019;26(2):303–5.

17. Matsuda K, Tanaka K, Fujishiro M, Saito Y, Ohtsuka K, Oda I, et al. Design paper: Japan Endoscopy Database (JED): A prospective, large database project related to gastroenterological endoscopy in Japan. Dig Endosc. 2018;30(1):5–19.

18. Matsuda T, Saika K. Cancer burden in Japan based on the latest cancer statistics: need for evidence-based cancer control programs. Annals of Cancer Epidemiology. 2018;2:2-.

19. Odagiri H, Yasunaga H, Matsui H, Fushimi K, Iizuka T, Kaise M. Hospital volume and the occurrence of bleeding and perforation after colorectal endoscopic submucosal dissection: analysis of a national administrative database in Japan. Dis Colon Rectum. 2015;58(6):597–603.

20. Yasunaga H, Horiguchi H, Kuwabara K, Matsuda S, Fushimi K, Hashimoto H, Ayanian JZ. Outcomes After Laparoscopic or Open Distal Gastrectomy for Early-Stage Gastric Cancer: A Propensity-Matched Analysis. Ann Surg. 2012.

21. Watanabe M, Ito H, Hosono S, Oze I, Ashida C, Tajima K, et al. Declining trends in prevalence of Helicobacter pylori infection by birth-year in a Japanese population. Cancer Sci. 2015;106(12):1738–43.

22. Inoue M. Changing epidemiology of Helicobacter pylori in Japan. Gastric Cancer. 2017;20(Suppl 1):3–7.

23. Takeuchi M, Endo H, Hibi T, Seishima R, Nakano Y, Yamamoto H, et al. The impact of COVID-19 for postoperative outcomes using a nationwide Japanese database of patients undergoing distal gastrectomy for gastric cancer. Ann Gastroenterol Surg. 2023;7(6):887–95.

24. Iijima K, Matsuhashi T, Shimodaira Y, Mikami T, Yoshimura T, Yanai S, et al. Impact of the COVID-19 pandemic on the performance of endoscopy in the Tohoku region of Japan. DEN Open. 2024;4(1):e249.

25. Japanese Gastric Cancer Association. Japanese Gastric Cancer Treatment Guidelines 2021 (6th edition). Gastric Cancer. 2023;26(1):1–25.

26. Soreide K, Sandvik OM, Soreide JA, Giljaca V, Jureckova A, Bulusu VR. Global epidemiology of gastrointestinal stromal tumours (GIST): A systematic review of population-based cohort studies. Cancer Epidemiol. 2016;40:39–46.

27. Ho V, Ku-Goto MH, Zhao H, Hoffman KE, Smith BD, Giordano SH. Regional differences in recommended cancer treatment for the elderly. BMC Health Serv Res. 2016;16:262.

28. Tanaka H, Ishikawa KB, Katanoda K. Geographic Access to Cancer Treatment in Japan: Results From a Combined Dataset of the Patient Survey and the Survey of Medical Institutions in 2011. J Epidemiol. 2018;28(11):470–5.

29. Watanabe T, Muro K, Ajioka Y, Hashiguchi Y, Ito Y, Saito Y, et al. Japanese Society for Cancer of the Colon and Rectum (JSCCR) guidelines 2016 for the treatment of colorectal cancer. Int J Clin Oncol. 2018;23(1):1–34.

30. Nobel TB, Lavery JA, Barbetta A, Gennarelli RL, Lidor AO, Jones DR, Molena D. National guidelines may reduce socioeconomic disparities in treatment selection for esophageal cancer. Dis Esophagus. 2019;32(5).

31. Ono H, Yao K, Fujishiro M, Oda I, Uedo N, Nimura S, et al. Guidelines for endoscopic submucosal dissection and endoscopic mucosal resection for early gastric cancer (second edition). Dig Endosc. 2021;33(1):4–20.

32. Bollschweiler E, Berlth F, Baltin C, Monig S, Holscher AH. Treatment of early gastric cancer in the Western World. World J Gastroenterol. 2014;20(19):5672–8.

